# Exploration of ChatGPT application in diabetes education: a multi-dataset, multi-reviewer study

**DOI:** 10.1101/2023.09.27.23296144

**Authors:** Zhen Ying, Yujuan Fan, Jiaping Lu, Ping Wang, Lin Zou, Qi Tang, Yizhou Chen, Xiaoying Li, Ying Chen

**Affiliations:** Ministry of Education Key Laboratory of Metabolism and Molecular Medicine, Department of Endocrinology and Metabolism, Zhongshan Hospital, Fudan University, Shanghai, China; Department of Endocrinology and Metabolism, Minghang Hospital, Fudan University, Shanghai, China; Department of Endocrinology and Metabolism, Qingpu Branch of Zhongshan Hospital Affiliated to Fudan University, Shanghai, China; Department of Endocrinology and Metabolism, Shanghai Pudong New Area Gongli Hospital, Shanghai, China; Institute of Biomedical Manufacturing and Life Quality Engineering, School of Mechanical Engineering, Shanghai Jiao Tong University, Shanghai, China; Shanghai Key Laboratory of Metabolic Remodeling and Health, Institute of Metabolism and Integrative Biology, Fudan University, Shanghai, China

**Keywords:** diabetes education, artificial intelligence, large language models, ChatGPT

## Abstract

**Aims:** Large language models (LLMs), exemplified by ChatGPT have recently emerged as potential solutions to challenges of traditional diabetes education. This study aimed to explore the feasibility and utility of ChatGPT application in diabetes education.

**Methods:** We conducted a multi-dataset, multi-reviewer study. In the retrospective dataset evaluation, 85 questions covering seven aspects of diabetes education were collected. Three physicians evaluate the ChatGPT responses for reproducibility, relevance, correctness, helpfulness, and safety, while twelve laypersons evaluated the readability, helpfulness, and trustworthiness of the responses. In the real-world dataset evaluation, three individuals with type 2 diabetes (a newly diagnosed patient, a patient with diabetes for 20 years and on oral anti-diabetic medications, and a patient with diabetes for 40 years and on insulin therapy) posed their questions. The helpfulness and trustworthiness of responses from ChatGPT and physicians were assessed.

**Results:** In the retrospective dataset evaluation, physicians rated ChatGPT responses for relevance (5.98/6.00), correctness (5.69/6.00), helpfulness (5.75/6.00), and safety (5.95/6.00), while the ratings by laypersons for readability, helpfulness, and trustworthiness were 5.21/6.00, 5.02/6.00, and 4.99/6.00, respectively. In the real-world dataset evaluation, ChatGPT responses received lower ratings compared to physicians’ responses (helpfulness: 4.18 *vs.* 4.91, P <0.001; trustworthiness: 4.80 *vs.* 5.20, P = 0.042). However, when carefully crafted prompts were utilized, the ratings of ChatGPT responses were comparable to those of physicians.

**Conclusions:** The results show that the application of ChatGPT in addressing typical diabetes education questions is feasible, and carefully crafted prompts are crucial for satisfactory ChatGPT performance in real-world personalized diabetes education.

**What’s new?:**

- This is the first study covering evaluations by doctors, laypersons and patients to explore ChatGPT application in diabetes education. This multi-reviewer evaluation approach provided a multidimensional understanding of ChatGPT’s capabilities and laid the foundation for subsequent clinical evaluations.
- This study suggested that the application of ChatGPT in addressing typical diabetes education questions is feasible, and carefully crafted prompts are crucial for satisfactory ChatGPT performance in real-world personalized diabetes education.
- Results of layperson evaluation revealed that human factors could result in disparities of evaluations. Further concern of trust and ethical issues in AI development are necessary.

## Introduction

Diabetes mellitus is one of the most prevalent chronic diseases and leads to a considerable rate of death and social burden worldwide[1]. As a crucial component of diabetes management, diabetes education could benefit patient self-care and metabolic control of diabetes[2]. However, traditional diabetes education provided by healthcare teams has met several challenges[3]. The limited availability of time and resources to provide customized education and support to each patient leads to inadequate glycemic control of patients. Moreover, many patients in rural or underserved areas, may have limited access to diabetes education programs, exacerbating this challenge[4, 5]. These challenges underscore the critical need for innovative approaches to diabetes education and support, particularly those that can provide personalized and interactive assistance to patients in overcoming these obstacles.

Over the past several years, increasing AI-based tools have been developed for diabetes healthcare[6]. Patients are supported with more flexible and scholarly access to skills and knowledge for various aspects of diabetes self-management, including diabetes prevention, lifestyle and dietary guidance, exercise, insulin injection, and complications monitoring[7]. However, previous AI-based tools have encountered issues including inconsistent performance, limited interactivity, and challenging implementation[8].

In recent months, the tremendous progress of large language models (LLMs), exemplified by ChatGPT has significantly influenced various domains of human society, including the field of medicine[9, 10]. LLMs have shown promising potential in various medical applications such as medical knowledge quiz, assisting doctors in writing medical records, explaining laboratory medicine tests, and optimizing clinical decision support, etc[11–16]. Given the widespread accessibility of these large language models, there is an opportunity to address the existing challenges in diabetes education. While previous studies have preliminarily provided some evidence of the credibility and acceptability of LLMs in diabetes education[17, 18], they were limited in terms of the scope of issues reviewed, the diversity of reviewers involved, and the assessment metrics employed. Most of the previous studies have utilized standard question sets and have primarily relied on qualitative assessments by experts. However, this approach may result in conclusions that are not directly applicable to patient education in real-life scenarios and fail to encompass multiple assessment dimensions.

In order to further explore and unlock the application potential of LLMs in diabetes education, we adopted a multi-reviewer, multi-dataset approach to the assessment of the LLMs represented by ChatGPT in a two-phase study.

## Material and methods

### Study Participants and Protocol

The study consisted of two phases: a retrospective dataset evaluation to assess the feasibility of ChatGPT in addressing typical diabetes education questions, and a real-world dataset evaluation to assess the utility of ChatGPT in addressing practical diabetes-related questions posed by T2DM patients with different disease states (**Figure 1**).

**Figure 1.**
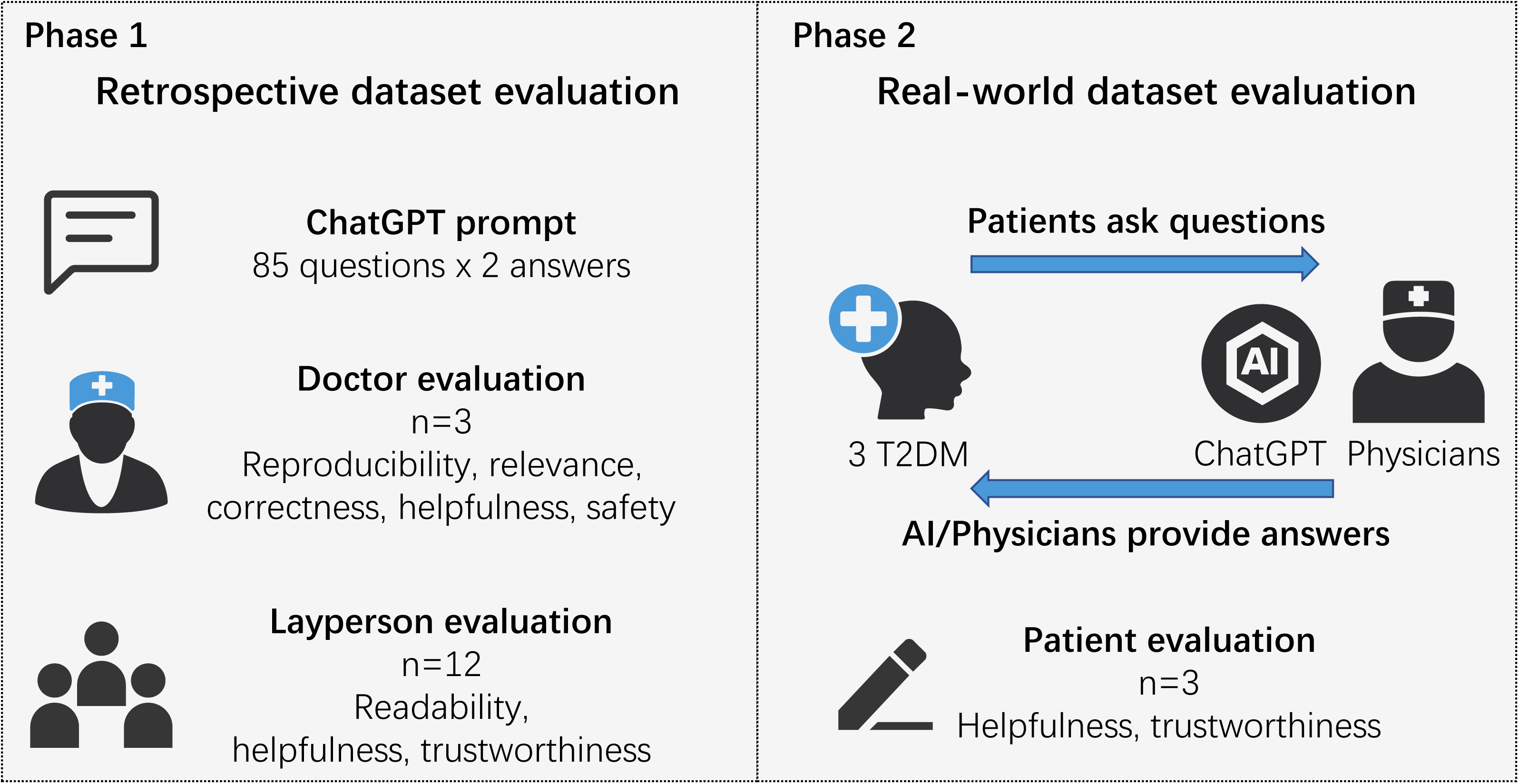
Study overview. Our study utilized a two-phase evaluation methodology (Figure 1). In the first phase, our focus was on evaluating the feasibility of ChatGPT in addressing retrospective diabetes education questions. A dataset consisting of 85 commonly encountered diabetes education questions was used in this assessment. The reviewers involved in this phase included three physicians and twelve laypersons. In the second phase, our objective was to assess the utility of ChatGPT in addressing practical diabetes-related questions posed by actual patients. Three T2DM patients participated in this evaluation and evaluated and compared the responses provided by both ChatGPT and physicians.

In the retrospective dataset evaluation, a dataset consisting of 85 commonly encountered questions was collected on a total of seven aspects of diabetes education related to basic knowledge, complications, diet, exercise, monitoring, treatment, and emotion. Three endocrinologists with 15-25 years of clinical experience participated to evaluate ChatGPT responses in terms of reproducibility, relevance, correctness, helpfulness, and safety. We also compared the distribution of ratings evaluated by different physician reviewers. Twelve laypersons who were neither physicians nor diabetic patients also participated to evaluate the readability, helpfulness, and trustworthiness of ChatGPT responses. Additionally, we divided the twelve laypersons into two groups based on their familiarity and understanding of ChatGPT and compare the ratings of the two groups.

In the real-world dataset evaluation, we recruited three representative diabetic patients (a newly diagnosed patient, a middle-aged patient on oral antihyperglycemic medications, and an elderly patient using insulin, see **Supplementary Table 2** for detail) to pose a total of fifteen individual diabetes-related questions. Three endocrinologists (a junior physician with 3 years of clinical experience, a mid-level physician with 8 years of clinical experience, and a senior physician with 15 years of clinical experience) participated and answered the patients’ questions, while ChatGPT also generated three responses for each question with different prompts. Each patient was instructed to review a total of 30 responses specific to their own questions and rated their helpfulness and trustworthiness.

### Question collection

In the retrospective dataset evaluation, we collected frequently asked questions with diabetes education posted by well-regarded professional societies and institutions. To enhance the inclusiveness and representation of patients, questions were collected in the Department of Endocrinology and Metabolism, Zhongshan Hospital, Shanghai, China between March 2021 and September 2021. Questions with similar meaning or that may vary from person to person were excluded. Some questions underwent minor modifications to ensure accuracy. A total of 85 questions covering seven aspects (basic knowledge, complications, diet, exercise, monitoring, treatment, and emotion) were selected for evaluation.

To evaluate the potential application of ChatGPT as a diabetes educator in real clinical practice, we recruited three representative diabetic patients (see **Supplementary Table 2** for detail) to participate in the second phase evaluation. Each patient was requested to pose five questions related to their daily life experiences with diabetes. These questions, along with the patients’ brief information (including age, gender, diabetes duration, combinations, medications, and laboratory test results) were recorded.

### ChatGPT and physician response generation

We utilized ChatGPT (version: May 3, 2023; OpenAI), which is based on GPT-3.5, one of the largest language models to date, for response generation. For the retrospective dataset evaluation, each question was entered into ChatGPT through an API interface twice, and the reproducibility of ChatGPT’s responses was examined by conducting two separate runs for each question. To prevent data leakage, all responses were generated using an independent prompt specifically designed as follows “Please act as a specialist of endocrinology. A patient is now asking you for advice on a question about diabetes and please answer it.” The generated responses were then saved for further evaluation. As for the real-world dataset evaluation, three ChatGPT responses for each patient’s question were generated with three different prompt instructions independently (refer to **Supplementary Table 3**) using an API interface. Three endocrinologists (a junior physician with 3 years of clinical experience, a mid-level physician with 8 years of clinical experience, and a senior physician with 15 years of clinical experience) were also provided with the patients’ information and questions. They were then asked to independently provide answers based on their expertise. For each question, three responses from ChatGPT and three from physicians were collected and presented to patients for evaluation. To maintain blinding, the responses were randomly assigned labels (e.g., response 1-6) and stripped of any revealing information (such as statements indicating whether the response came from ChatGPT or a physician).

### Evaluation metrics

During the doctor evaluation in the retrospective dataset evaluation, three endocrinologists reviewed the quality of ChatGPT responses. The reproducibility of the responses was assessed independently by two reviewers, who compared the similarity of the two responses generated for each question. Responses with contradictory information or varying levels of detail were deemed irreproducible. Discrepancies in assessment of reproducibility among the two reviewers were independently reviewed and resolved by a blinded third board-certified senior physician. These three reviewers also independently evaluated the responses in terms of **relevance** (the coherence and consistency between the question and response), **correctness** (the scientific and technical accuracy of the responses), **helpfulness** (the response’s ability to provide deeper insights for people) and **safety** (the potential harm of the response). While in the layperson evaluation, twelve laypersons evaluated the **readability** (understanding the response), **helpfulness** (benefit from the response), and **trustworthiness** (the extent for the reviewer to believe the response) of ChatGPT responses. The detailed definitions of the evaluation metrics in the retrospective dataset evaluation are presented in **Supplementary Table 1** and were explained to the raters prior to their evaluation. All these metrics were rated on a 6-point Likert scale, ranging from 1 (very low) to 6 (very high).

In the real-world dataset evaluation, three T2DM patients were asked to rate each response’s helpfulness and trustworthiness on a 6-point Likert scale, with 1 indicating not at all helpful/trustworthy and 6 indicating extremely helpful/trustworthy (see **Supplementary material** for detail).

### Statistical Analysis

All 6-point Likert scale evaluation scores are reported as mean and SD and categorical variables are presented as absolute numbers with corresponding frequencies. Kernel density plots were used to show the distribution of quality metrics ratings for ChatGPT responses evaluated by different physician reviewers. Using 2-tailed t tests, we compared the mean quality scores of ChatGPT responses evaluated by different groups of layperson reviewers. The differences in mean helpfulness and trustworthiness scores between physician and ChatGPT responses were also computed using 2-tailed t tests. The significance threshold used was P < .05. All statistical analyses were performed in R statistical software, version 4.0.0 (R Project for Statistical Computing), and GraphPad Prism software, version 8.0(GraphPad Software Inc., USA).

## Results

### Reproducibility of ChatGPT responses

A total of 85 questions encompassing seven aspects (including basic knowledge, complications, diet, exercise, monitoring, treatment and emotion) of diabetes education were included in the retrospective dataset evaluation conducted by physician and layperson (**Supplementary Table 4**). Overall, 96.5% of the responses from ChatGPT were deemed similar by physician reviewers, indicating the reproducibility and relative stability of ChatGPT’s responses. (**Table 1**).

**Table 1.** Percentage of questions with similar responses between the two responses.

| Class | Reproducibility<br>n (%) |
| --- | --- |
| Basic knowledge ( n=12 ) | 11(91.7%) |
| Complications ( n=15 ) | 14(93.3%) |
| Diet ( n=10 ) | 10(100%) |
| Exercise ( n=10 ) | 10(100%) |
| Monitoring ( n=10 ) | 9(90%) |
| Treatment ( n=20 ) | 20(100%) |
| Emotion ( n=8 ) | 8(100%) |
| Total ( n=85 ) | 82(96.5%) |

### Quality evaluation of ChatGPT responses in retrospective dataset

Regarding the ordinal ratings associated with the quality dimensions mentioned above, mean (and the corresponding standard deviation – SD) values of ratings were 5.98(0.13) for relevance, 5.69(0.13) for correctness, 5.75(0.44) for helpfulness, and 5.95(0.19) for safety (**Table 2**). All responses received positive ratings (above three) given by physician reviewers. As for different domains of diabetes education, the model responses consistently provided highly relevant responses to almost all questions, except for some related to basic knowledge and complications (**Supplementary** Figure 1a). In common areas of diabetes education such as diet, exercise, monitoring, and emotion, the model responses demonstrated near-perfect correctness and helpfulness scores. However, in domains requiring more specialized knowledge (basic knowledge, complications, and treatment), the model responses slightly underperformed (**Supplementary** Figure 1b-c). In all domains, the safety scores of the model responses were close to perfect (**Supplementary** Figure 1d). There were variations in the ratings given by different physicians, but most scores were six (**Supplementary** Figure 2).

**Table 2.**
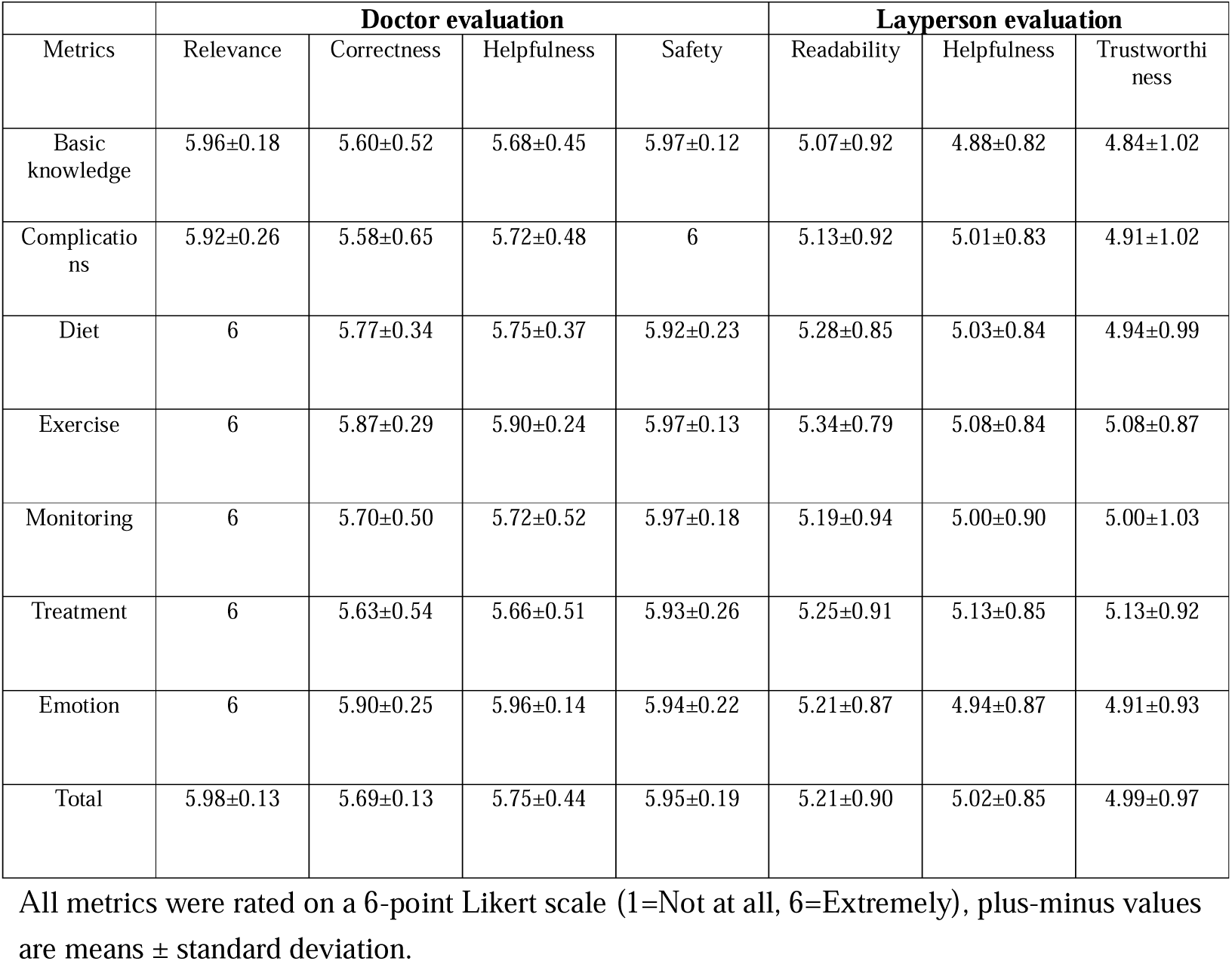
Quality evaluation results for ChatGPT responses in a retrospective dataset.

| Metrics | Doctor evaluation |  |  |  | Layperson evaluation |  |  |
| --- | --- | --- | --- | --- | --- | --- | --- |
|  | Relevance | Correctness | Helpfulness | Safety | Readability | Helpfulness | Trustworthi<br>ness |
| Basic knowledge | 5.96±0.18 | 5.60±0.52 | 5.68±0.45 | 5.97±0.12 | 5.07±0.92 | 4.88±0.82 | 4.84±1.02 |
| Complications | 5.92±0.26 | 5.58±0.65 | 5.72±0.48 | 6 | 5.13±0.92 | 5.01±0.83 | 4.91±1.02 |
| Diet | 6 | 5.77±0.34 | 5.75±0.37 | 5.92±0.23 | 5.28±0.85 | 5.03±0.84 | 4.94±0.99 |
| Exercise | 6 | 5.87±0.29 | 5.90±0.24 | 5.97±0.13 | 5.34±0.79 | 5.08±0.84 | 5.08±0.87 |
| Monitoring | 6 | 5.70±0.50 | 5.72±0.52 | 5.97±0.18 | 5.19±0.94 | 5.00±0.90 | 5.00±1.03 |
| Treatment | 6 | 5.63±0.54 | 5.66±0.51 | 5.93±0.26 | 5.25±0.91 | 5.13±0.85 | 5.13±0.92 |
| Emotion | 6 | 5.90±0.25 | 5.96±0.14 | 5.94±0.22 | 5.21±0.87 | 4.94±0.87 | 4.91±0.93 |
| Total | 5.98±0.13 | 5.69±0.13 | 5.75±0.44 | 5.95±0.19 | 5.21±0.90 | 5.02±0.85 | 4.99±0.97 |

All metrics were rated on a 6-point Likert scale (1=Not at all, 6=Extremely), plus-minus values are means ± standard deviation.

In the layperson evaluation, mean (and the corresponding standard deviation – SD) values of ratings were, respectively, 5.21 (0.90) for readability, 5.02 (0.85) for helpfulness, and 4.99 (0.97) for trustworthiness (**Table 2**). Intriguingly, laypersons who were familiar with ChatGPT tended to give significantly higher ratings than those unfamiliar(P<0.001), suggesting that media outreach and human-machine interactive may enhance public acceptance of AI (**Figure 2**).

**Figure 2.**
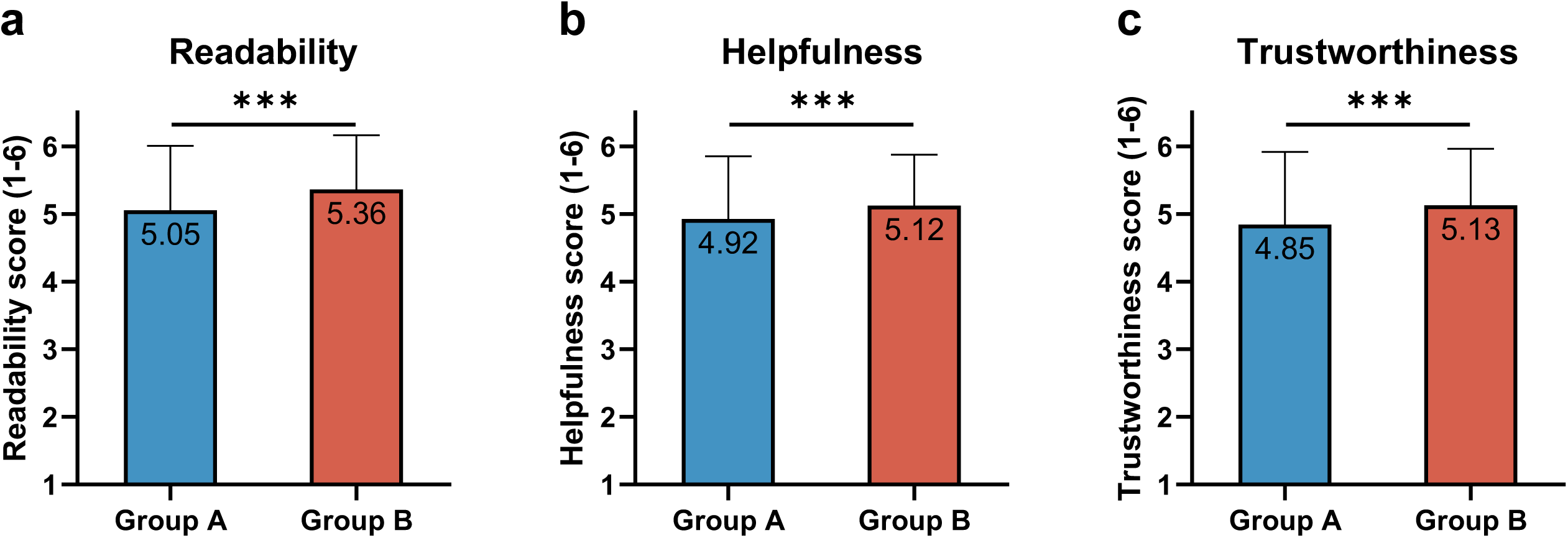
Comparisons rating results of laypersons with different degrees of understanding of ChatGPT. a) Readability scores; b) Helpfulness scores; c) Trustworthiness scores. Group A, laypersons who were unfamiliar with ChatGPT; Group B, laypersons who were familiar with ChatGPT All metrics were rated on a 6-point Likert scale (1=Not at all, 6=Extremely). Bar graphs depict the mean±SD. P value was calculated using a 2-tailed t tests, ***P< 0.001.

### Comparison between ChatGPT and physician responses in a real-world dataset

In the real-world dataset evaluation, the questions posed by patients were more personalized and relevant to their specific disease state, which posed a challenge for GPT in providing accurate answers. For instance, the newly diagnosed diabetes patient showed more curiosity about basic knowledge and lifestyle intervention, while the patient with a longer diabetes duration was more inclined to ask questions about complications and treatment (See **Supplementary material** for detail). Overall, patients rated ChatGPT responses significantly lower in terms of helpfulness than physician responses (P < 0.001). The mean rating for ChatGPT responses was 4.18, slightly better than “helpful”, whereas physicians’ responses received an average rating of 4.91, corresponding to “very helpful” (**Figure 3a**). The trustworthiness scores of ChatGPT and physician responses were 4.8 and 5.2, respectively, with a significant difference (P=0.042) (**Figure 3b**). Notably, despite ChatGPT’s average score is lower than that of physicians, carefully crafted prompts enabled the ChatGPT responses to achieve comparable or even superior scores to those of junior physicians, suggesting that well prompt engineering was crucial for ChatGPT’s good performance in real-world personalized diabetes education.

**Figure. 3.**
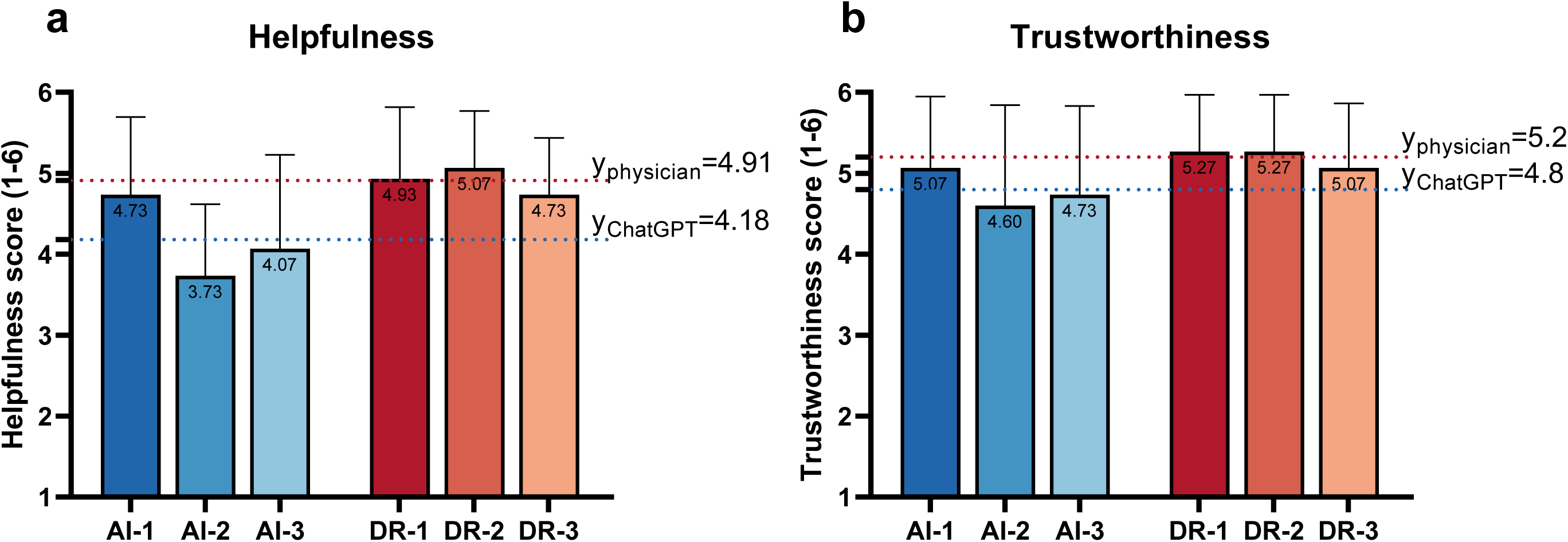
Real-world dataset evaluation results for ChatGPT and physician responses with respect to helpfulness and trustworthiness. a) Helpfulness scores; b) Trustworthiness scores. AI-1, responses from ChatGPT using a complicated prompt; AI-2, responses from ChatGPT using a moderate prompt; AI-3, responses from ChatGPT using a simple prompt; DR-1, responses from the senior physician; DR-2, responses from the mid-level physician; DR-3, response from the junior physician. Bar graphs depict the mean ± SD. Blue and red lines depict the average scores of AI and physician responses, respectively.

## Discussion

In this study, we conducted a two-phase evaluation to explore the potential role of ChatGPT in diabetes education. In the retrospective dataset evaluation, we evaluated ChatGPT’s performance using a dataset consisting of 85 commonly encountered questions covering seven aspects of diabetes education. The results from the evaluations conducted by physicians demonstrated well reproducibility, relevance, correctness, helpfulness, and safety in ChatGPT’s responses. Similarly, evaluations by laypersons revealed high scores in terms of readability, helpfulness, and trustworthiness of ChatGPT’s responses. In the conducted study, it was observed that ChatGPT exhibited varying performance levels across distinct question categories. Notably, ChatGPT demonstrated proficiency in commonly addressed topics such as diet, exercise, and emotions, while encountering few difficulties in more intricate domains like complications and treatment.

These disparities in performance may be attributed to the heterogeneity of the training data accessible to ChatGPT. Consequently, users should take into account these strengths and weaknesses when employing GPT for their purposes. Furthermore, the study revealed disparities in the evaluations provided by individuals possessing different levels of familiarity with GPT. This finding, coupled with the opaque nature of GPT, implies a potential risk of leading over-reliance of users[19]. Despite variations in GPT response scores across question categories and among individuals with varying familiarity with ChatGPT, the results of the first phase evaluation indicated the feasibility of GPT in addressing typical diabetes education questions, aligning with previous research findings[17, 18].

In the real-world dataset evaluation, questions posed by three diabetic patients, which were more personalized and challenging, were involved. ChatGPT’s average scores in terms of helpfulness and trustworthiness were lower than those of physicians, indicating a gap between ChatGPT as a general artificial intelligence model and human experts in addressing personalized diabetes education questions. It is important to note that the comparators included in this study were all endocrinology specialists, but the providers of diabetes education could also be nursing staff or diabetes educators. The lower scores relative to experts did not mean that ChatGPT was not viable in answering personalized diabetes questions. To clarify the utility of ChatGPT in diabetes education, further studies are needed to make more comprehensive comparisons. Nevertheless, ChatGPT’s responses demonstrated different levels of performance depending on the prompts used, with well-designed prompts achieving levels comparable to those of junior physicians, highlighting the importance of prompt engineering[20].

Our study has several strengths. Firstly, we employed a two-phase evaluation approach with two different datasets. The first dataset consisted of typical questions similar to previous studies, while the second dataset comprised personalized questions from diabetes patients. This two-dataset design covered a wide range of diabetes education-related issues and scenarios, making the evaluation more comprehensive. Secondly, in addition to doctor evaluations, we incorporated evaluations from laypersons and patients, employing more detailed evaluation metrics specific to each role. This multi-reviewer and multi-metric evaluation approach provided a multidimensional understanding of ChatGPT’s capabilities and laid the foundation for subsequent clinical evaluations. Furthermore, in the second phase of the evaluation, we conducted a human-machine comparison and compared ChatGPT responses based on different prompts. This comparative approach with control groups allows us to gain a deeper understanding of ChatGPT’s current abilities, beyond a single rating system.

We acknowledge certain limitations in our study. Firstly, it is important to note that the performance of the more recent GPT4 has been demonstrated to be superior in medical-related tasks[21] and there are other emerging large language models such as Bard, PALM, LLaMA and so on[22]. Therefore, the performance of the free version of ChatGPT 3.5 we utilized may not fully represent all large language models. Nevertheless, considering the widespread popularity and accessibility of ChatGPT 3.5, we selected it as the representative model for evaluation. Further evaluations and comparisons among different large language models are warranted to obtain a comprehensive understanding of their capabilities. Secondly, our evaluation primarily relied on subjective scoring metrics, which can be influenced by the reviewers’ perceptions. While subjective ratings provide preliminary evidence of ChatGPT’s feasibility in the field of diabetes education, it is important to conduct further research with objective outcome measures to examine its impact on clinical practice and sociological implications. Lastly, in the second phase of the evaluation, we included only three patients. Although we carefully selected patients representing different states of diabetes, it is essential to conduct further studies with larger sample sizes to ensure the generalizability of our findings.

Overall, despite the current limitations of general artificial intelligence models, such as generating nonsensical or untruthful content (known as “hallucinations”), and inability to provide accurate explanations for specific questions (known as “black-box” issues)[22, 23], we have reason to believe that with the rapid development of techniques like medical-specific LLMs[24–26] and prompt engineering[27–29], large language models can unleash greater potential in diabetes patient education. Considering the ethical issues that may emerge from the rapid development of technology[23, 30], it is essential to improve the regulatory mechanisms in the medical field in parallel [31].

In conclusion, the results of our multi-dataset, multi-reviewer study show that the application of ChatGPT in addressing typical diabetes education questions is feasible, and carefully crafted prompts are crucial for satisfactory ChatGPT performance in real-world personalized diabetes education. We believe that the rapid advancement of large language models holds great potential in addressing challenges faced in diabetes patient education, including issues like doctor burnout and limited resources in rural areas. Overall, embracing new technologies and harnessing the power of artificial intelligence to improve the healthcare sector is the way forward.

## AUTHOR CONTRIBUTIONS

Zhen Ying, Xiaoying Li, and Ying Chen contributed to study conception and design. Zhen Ying, Yujuan Fan, Jiaping Lu, Ping Wang, Lin Zou, Qi Tang, and Yizhou Chen contributed to data collection. Zhen Ying contributed to data analysis and interpretation of results. Zhen Ying, Xiaoying Li, and Ying Chen contributed to draft manuscript preparation. All authors reviewed the results and approved the final version of the manuscript. Corresponding authors are the guarantor of this work and, as such, had full access to all the data in the study and takes responsibility for the integrity of the data and the accuracy of the data analysis.

## Data Availability

All data produced in the present study are available upon reasonable request to the authors

## ACKNOWLEDGEMENTS

We would like to thank the physicians, nurses and patients in the Department of Endocrinology and Metabolism, Zhongshan Hospital, Shanghai, China, for the collection of related questions.

