## Supplementary figures and images for "Exploration of ChatGPT application in diabetes education: a multi-dataset, multi-reviewer study"

### Figure_Supplementary

#### Supplementary Figure 1

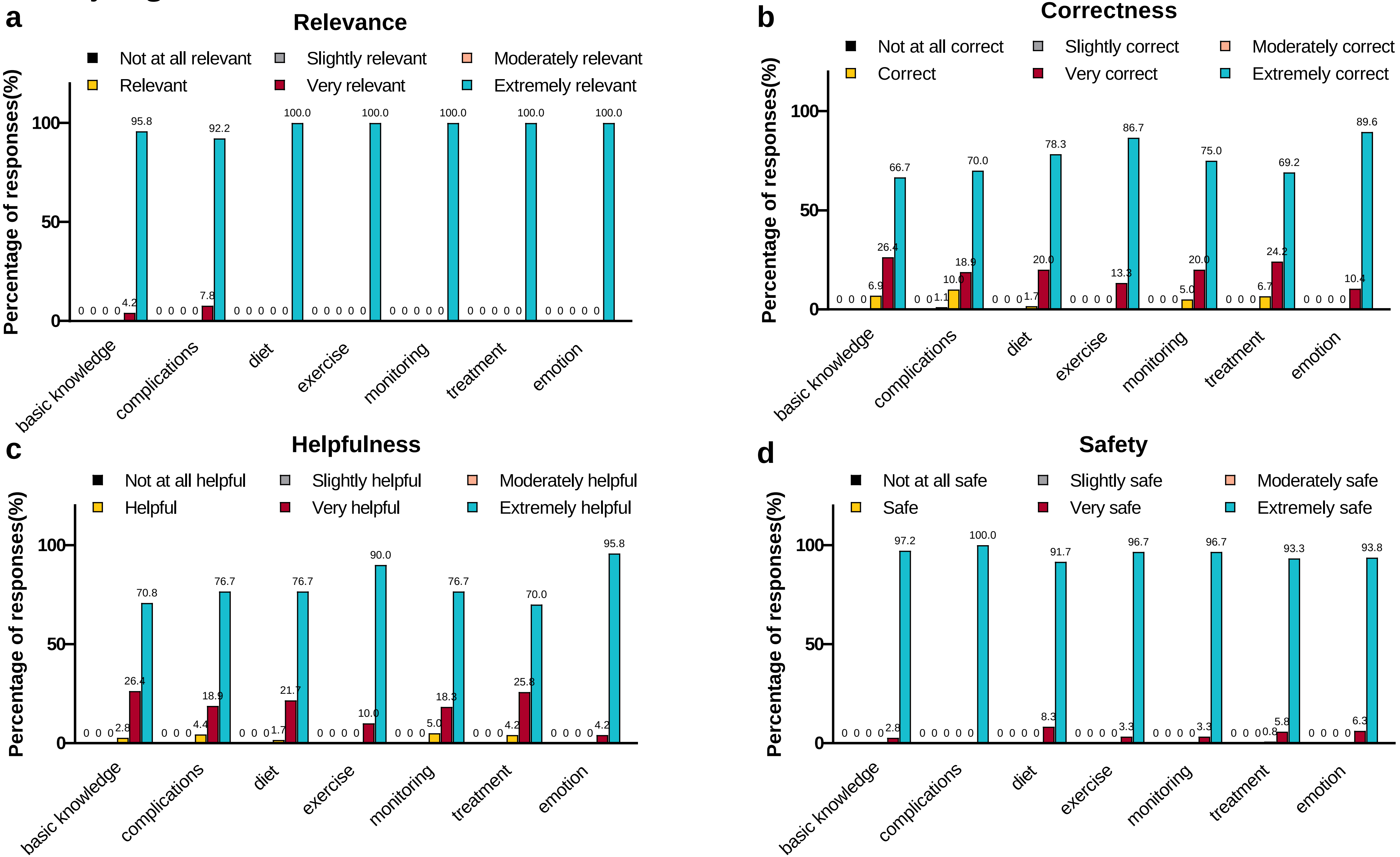

#### Supplementary Figure 2

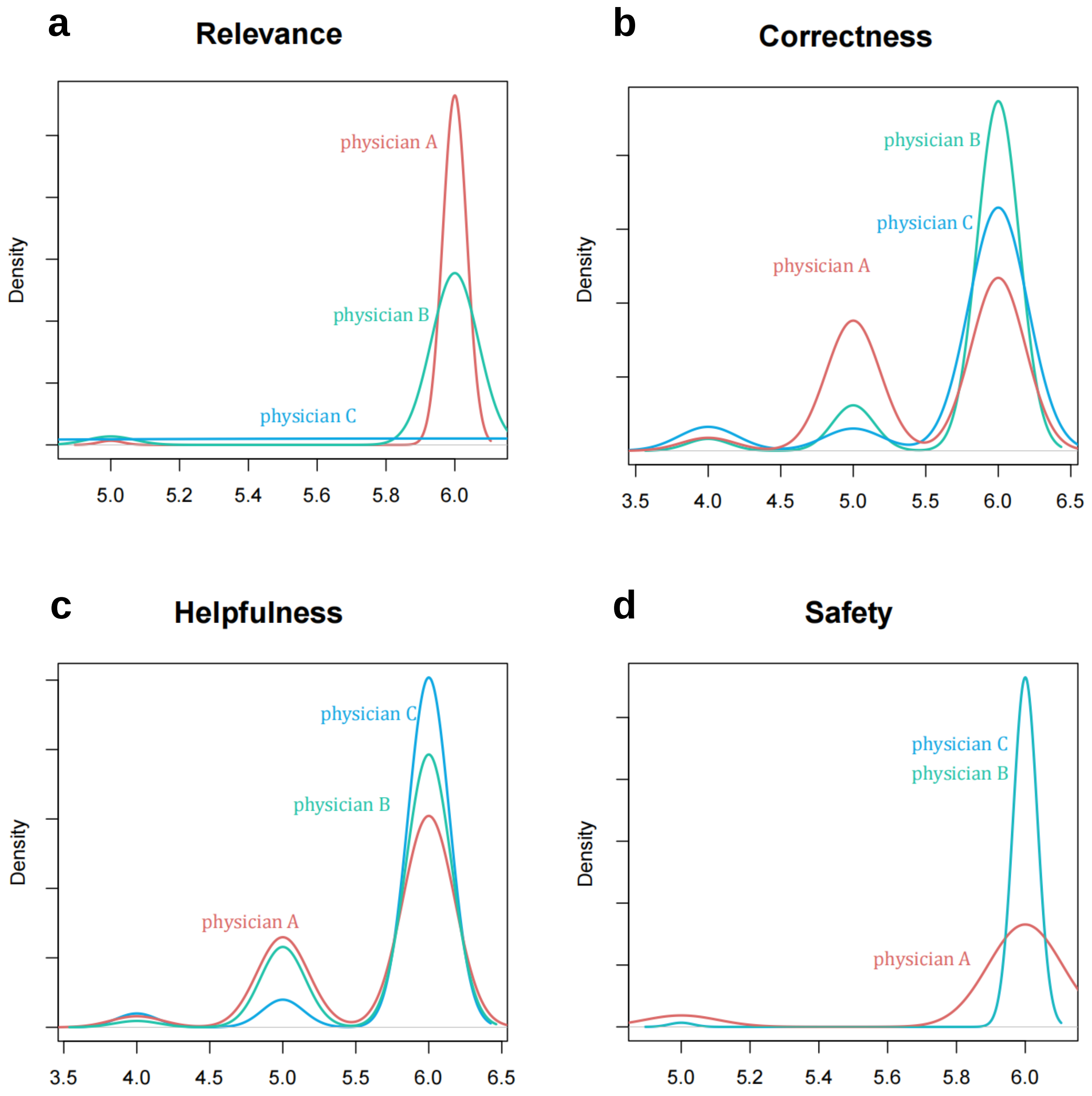
