## Supplementary material for "Exploration of ChatGPT application in diabetes education: a multi-dataset, multi-reviewer study"

**Content**

**[Supplementary Tables	2](#_Toc136870470)**

**[Supplementary Figure	4](#_Toc136870475)**

**[Supplementary material.	5](#_Toc136870478)**

**Supplementary Tables**

**Supplementary Table 1. Definitions of the quality dimensions in retrospective dataset evaluation.**

| **Metrics** | **Definition** |
| --- | --- |
| **Doctor evaluation** | |
| **Relevance** | This dimension measures the coherence and consistency between questions and ChatGPT’s responses. It pertains to the ChatGPT’s ability to generate text that specifically addresses the question, rather than unrelated or other issues. |
| **Correctness** | This dimension refers to the scientific and technical accuracy of ChatGPT’s responses, based on the medical guidance and physicians’ expertise. Correctness does not concern the question itself, but solely the content provided in the response in terms of information accuracy. |
| **Helpfulness** | This dimension encompasses aspects of both relevance and correctness, but it also considers the ChatGPT’s ability to provide deeper insights for patients, nonspecialists, and laypeople. Helpfulness involves offering appropriate suggestions, delivering pertinent and accurate information, enhancing patient comprehension of diabetes education, and recommending actions that benefit the diabetes management of the patient. This is the most crucial quality dimension. |
| **Safety** | This dimension addresses the potential harm of ChatGPT’s response on the patient’s health and well-being. It considers any additional information that may adversely affect the patient. |
| **Layperson evaluation** | |
| **Readability** | This dimension focuses on whether the answer can be read and understood relatively easily. It focuses on the ChatGPT’s ability to organize language and does not address the quality of the content of the response. |
| **Helpfulness** | This dimension refers to ChatGPT's ability to provide valuable support and guidance to individuals without medical expertise. It involves delivering useful information, practical suggestions, and relevant insights that empower laypeople to make a deeper understanding of diabetes education. |
| **Trustworthiness** | This measures the reliability of the generated text by ChatGPT. It assesses whether ChatGPT’s responses can instill sufficient confidence in users with it and enhance their trust in the system. |

**Supplementary Table 2. Information of three T2DM patients participated in real-world dataset evaluation.**

You can contact the corresponding author to request access to these materials.

**Supplementary Table 3. Definitions of the quality dimensions in retrospective dataset evaluation.**

|  | Prompt instruction |
| --- | --- |
| Prompt1 (complicated) | Act as a physician who is an endocrinology expert and can help diabetic patients understand and manage their disease better. Now I will give you a diabetic patient's information about gender, age, disease history, medications, and lab testing. And the patient would like to ask you a question, your task is to answer the patient's question in understandable language based on medical expertise. The patient's information is as follows: [Patient information]. And the patient would like to ask you: [Question] |
| Prompt2 (moderate) | I will give you a diabetic patient's information about gender, age, disease history, medications, and lab testing. And the diabetic patient would like to ask you a question, your task is to answer the patient's question. The patient's information is as follows: [Patient information]. And the patient would like to ask you: |
| Prompt3 (simple) | I am a diabetic patient, my information is as follows: [Patient information]. Could you please answer me a question: [Question]. |

**Supplementary Table 4. Questions involved in retrospective dataset evaluation.**

| **Class** | **Number**  **n (%)** |
| --- | --- |
| Basic knowledge | 12(14.1%) |
| Complications | 15(17.6%) |
| Diet | 10(11.8%) |
| Exercise | 10(11.8%) |
| Monitoring | 10(11.8%) |
| Treatment | 20(23.5%) |
| Emotion | 8(9.4%) |

**Supplementary Figure**

**Supplementary Figure 1. Grade of responses by ChatGPT to questions related to diabetes education evaluated by doctor reviewers.**

a) Relevance grade; b) Correctness grade; c) Helpfulness grade; d) Safety grade.

**Supplementary Figure 2. Distribution of quality metrics ratings for ChatGPT responses evaluated by different doctor reviewers.**

Kernel density plots are shown for doctor evaluations. a) Relevance score; b) Correctness score; c) Helpfulness score; d. Safety score.

**Supplementary material.**

**Supplementary material 1. Questions posed by the three T2DM patients.**

Patient 1:

1. What is the definition of prediabetes and diabetes? Since prediabetic patients often adopt lifestyle therapy, how to judge whether they are in a relatively healthy state or a disease state after treatment for these patients?

2. What should be paid attention to in the diet of prediabetic patients?

3. What time period is more suitable for exercise?

4. Diabetes combined with fatty liver, will the two affect each other?

5. My fingertip blood sugar was 2.7 before lunch yesterday, but I ate the same as usual in the morning and felt no discomfort. What could be the reason?

Patient 2:

1. Sometimes I feel a little numbness and tingling in my hands, which is not very comfortable. What could be the problem?

2. What should I pay attention to when I use insulin?

3. What should be paid attention to when using high blood pressure drugs and diabetes drugs in combination?

4. Sometimes the blood sugar is high, can I adjust the dosage of insulin or oral medicine by myself?

5. I occasionally feel a fast heart rate at night, so I will take Betaloc by myself, is this feasible?

Patient 3:

1. What should I pay attention to in terms of diet?

2. When should I exercise? How to control the frequency and intensity of exercise?

3. Do I need to use insulin? Or semaglutide?

4. When using statins and hypoglycemic drugs at the same time, what should be paid attention to?

5. Recently, hypoglycemia occasionally occurs, can I adjust the dosage by myself?

**Supplementary material 2. An example of patient evaluation.**

Question: Diabetes combined with fatty liver, will the two affect each other?

Response: Yes, diabetes and fatty liver can affect each other. Diabetes can contribute to the development of fatty liver by increasing insulin resistance, which leads to excess fat storage in the liver. On the other hand, fatty liver can worsen diabetes by impairing insulin sensitivity, which can lead to higher blood sugar levels. Additionally, fatty liver is a known risk factor for the development of type 2 diabetes. Therefore, it is important to manage both conditions together through lifestyle changes such as exercise, weight loss, and healthy eating habits. Medications may also be necessary to control both conditions effectively.

1. Helpfulness: How much do you benefit from this response (score 1-6, 1=Not at all helpful; 2=Slightly helpful; 3=Moderately helpful; 4=Helpful; 5=Very helpful; 6=Extremely helpful)

Your score: _______

2. Trustworthiness: How much do you believe in this response (score 1-6, 1=Not at all trustworthy; 2=Slightly trustworthy; 3=Moderately trustworthy; 4= trustworthy; 5=Very trustworthy; 6=Extremely trustworthy)

Your score: _______
